# Reverse Transcription-Loop-Mediated Isothermal Amplification (RT-LAMP) is an effective alternative for SARS-CoV-2 molecular detection in middle-income countries

**DOI:** 10.1101/2020.10.14.20212977

**Authors:** Oscar Escalante-Maldonado, Margot Vidal-Anzardo, Fernando Donaires, Gilmer Solis-Sanchez, Italo Gallesi, Luis Pampa-Espinoza, Maribel Huaringa, Nancy Rojas Serrano, Coralith García, Eddie Angles-Yanqui, Ronnie Gustavo Gavilán, Ricardo Durães-Carvalho, Cesar Cabezas, Paulo Vitor Marques Simas

**Affiliations:** Instituto Nacional de Salud, Lima, Peru; Hospital Nacional Cayetano Heredia, Lima, Peru; Hospital Nacional Arzobispo Loayza, Lima, Peru; Universidad Peruana Cayetano Heredia, Lima, Peru; Universidad Nacional Mayor de San Marcos, Lima, Peru; University of Campinas, Institute of Biology, Laboratory of Animal Virology, Campinas, SP, Brazil

**Keywords:** COVID-19, molecular testing, RT-LAMP, healthcare unit

## Abstract

Molecular diagnosis of SARS-CoV-2 in developing countries is still a big challenge. The reference standard, RT-qPCR, recommended by WHO, is not widely available, difficulting early identification of cases. Furthermore, the transport logistic between the sample collection point and the laboratory facilities can alter the samples, producing false negative results. RT-LAMP is a cheaper, simpler molecular technique that can be an interesting alternative to be offered in hospital laboratories. We present the evaluation of a RT-LAMP for diagnosis of SARS-CoV-2 in two steps: the laboratory standardization and the clinical validation, comparing it with the standard RT-qPCR. In the standardization phase, limit of detection and robustness values were obtained using RNA from a Peruvian SARS-CoV-2 strain. It presented 100% agreement between triplicates (RT-LAMP agreement with all RT-qPCR reactions that presented Ct ≤ 30) and robustness (RT-LAMP successful reactions with 80% reaction volume and 50% primer concentration). 384 nasal and pharyngeal swabs collected from symptomatic patients and stored in the INS biobank were tested and we obtained 98.75%, 87.41%, 97.65% and 92.96% for specificity, sensitivity, positive predictive value and negative predictive values respectively. Then, 383 samples from symptomatic patients with less than 15 days of disease, were tested both with the RT-LAMP and with the RT-qPCR, obtaining e 98.8%, 88.1%, 97.7% y 93.3% of specificity, sensitivity, positive predictive value and negative predictive values respectively. The laboratory standardization and the clinical validation presented the same value by Kappa-Cohen index (0.88) indicating an almost perfect agreement between RT-LAMP and RT-qPCR for molecular detection of SARS-CoV-2. We conclude that this RT-LAMP protocol presented high diagnostic performance values and can be an effective alternative for COVID-19 molecular diagnosis in hospitals, contributing to early diagnosis and reducing the spread of virus transmission in the Peruvian population.

## 1. INTRODUCTION

The World Health Organization (WHO) declared COVID-19 as a pandemic in the beginning of March. Since, the virus has been detected in every continent and produced more than 1 million deaths. Currently, some Latin America countries such as Brazil and Peru are considered pandemic epicenters [1], but many more low and middle –income countries are facing important health constraints.

Molecular tests require considerable financial and logistical investments, when compared to other diagnostic tools. The reference standard test suggested by WHO, the Real Time Reverse Transcription Polymerase Chain Reaction (RT-qPCR), requires molecular laboratory facilities, uses expensive equipment (thermocycler), reagents (probes) and specialized staff all of which are not always widely available in these countries. Results are available between 4 and 8 hours of processing [2, 3].

In Peru, at the beginning of the pandemic, RT-qPCR was only able to be performed in a standardized way in Lima (capital of the country) in the National Reference Laboratory of Respiratory Viruses of the Instituto Nacional de Salud (INS). Progressively, the diagnosis was extended to regional laboratories in a decentralized manner, but the demand for these tests, in practice, has not been fully met in some places. This situation has led to the concern of the local scientific community for the development of diagnostic alternatives.

On the other hand, the simple and low-cost reverse transcription loop-mediated isothermal amplification (RT-LAMP) method could be a good alternative for molecular diagnosis in places where there is no complete laboratory infrastructure, particularly in hospitals. It is an isothermal technique that uses from four to six primers, two/three forward and two/three reverse to identify DNA targets to allow its amplifications. RT-LAMP uses cheaper equipment, is fast (results generally available in almost 50 minutes, without considering sampling and RNA extraction time) and highly sensible [4]. There are several publications about this technique, showing good results when compared to the RT-qPCR method.

Our goal was to develop a RT-LAMP for molecular detection of SARS-CoV-2 and to evaluate its diagnostic performance both through basic laboratory standardization as well as through assessment of diagnostic parameters in patients with clinical suspicion of COVID-19, comparing it with RT-qPCR as the reference standard.

## 2. MATERIAL AND METHODS

### 2.1. ETHICAL CONSIDERATIONS

The laboratory standardization did not need to be sent for evaluation by the Ethics Committee since it is included in the action plan of INS-Peru. Nonetheless, all samples were processed completely anonymously. The clinical validation protocol was submitted to the Ethics Committee of the INS-Peru and approved on August 6^th^, 2020, under the procedure “Revisión de protocolos en el marco de epidemias, brotes o situaciones de emergencia” as indicates RD No. 283-2020-OGITT-INS.

### 2.2. EXPERIMENTAL DESIGN

The diagnostic performance values of RT-LAMP in comparison to RT-qPCR were obtained from qualitative and quantitative parameters used for laboratory standardization and clinical assessment. All experiments were conducted under the same conditions (samples, equipment, technicians and environment).

### 2.3. SAMPLES AND EVALUATION PARAMETERS

#### 2.3.1. LABORATORY STANDARDIZATION

The limit of detection and the robustness (concordance degree of the results when we change primers concentration – 0.5P – and the final volume of reaction– 0.8V, 0.6V, 0.5V and 0.4V) were performed using a SARS-CoV-2 Peruvian strain isolated and titred in Vero cell line. The cross-reaction analysis was performed *in silico* using multiple sequences alignment between external primers of RT-LAMP and reference sequences for all known human coronaviruses (HCoV) (NC_005831.2, HCoV-NL63; NC_002645.1, HCoV-229E; NC_006213.1, HCoV-OC43 strain ATCC VR-759; NC_006577.2, HCoV-HKU1; NC_004718.3, Severe Acute Respiratory Syndrome-related Coronavirus Type 1; NC_019843.3, Middle East Respiratory Syndrome-related Coronavirus; FJ415324.1, HECoV 4408) and SARS-CoV-2 strains from strains from China (NC_045512.2) and Peru (all complete sequences made available on the GISAID) [5].

Specificity, sensitivity positive and negative predictive values were obtained through evaluation of 384 nasal and pharyngeal swabs collected from routine epidemiological screening. From these, 193 were submitted to a new RT-LAMP round by other laboratory technician and equipment to test the reproducibility. The sample size was calculated using the formula for difference between 2 proportions assuming a 90% power and a 95% confidence interval [6] from the total number of samples processed by RT-qPCR (almost 240,000 samples until July 2020).

#### 2.3.2. CLINICAL ASSESSMENT

Specificity, sensitivity, positive and negative predictive values were obtained through evaluation of 383 COVID-19 suspected people up to 15 days after symptom onset, from Lima, Peru, assessed in Hospital Cayetano Heredia, Hospital Hipolito Unanue and Hospital Arzobispo Loayza and patients that were treated by home care teams. The sample size was calculated using Epidat software version 4.2, considering an estimate of 91.489% sensitivity and 99.531% specificity according to Jiang et al. [7]. People older than 18 years old without a previous diagnosis of COVID-19 by molecular test were included in the study after signature of informed consent. Pregnant women and severe or critical patients were excluded. The validation criteria considered 95% significance level, 5% absolute error and 39.5% positivity probability (based in the positive results obtained by RT-qPCR reported by INS-Peru and assuming a loss rate of 20%). Nasal and pharyngeal swabs were performed on each subject, using the Yocon Biology Technology Company sampling kit, which includes viral transport media and flocked dacron swabs. The samples were transported to the INS-Peru using triple containers with cold accumulators, at temperatures between 2 to 8 ° C.

### 2.4. MOLECULAR DETECTION OF SARS-CoV-2

#### 2.4.1. RNA EXTRACTION

The RNA extraction was performed using GenElute™ Total RNA Purification Kit (Sigma-Aldrich – Merck), according to manufacturers’ instructions, then quantified by NanoDrop™ Spectrophotometer and frozen to −80°C until further processing.

#### 2.4.2. RT-qPCR REACTION

The primers and probes used in the RT-qPCR reactions standardized by INS-PERU, is available in table 1. The RT-qPCR was performed using Rotor-Gene Multiplex RT-PCR Kit, according to the RT-qPCR standardized and implemented to COVID-19 diagnosis at the INS-Peru, summarized in the tables 2 (reactions conditions) and 3 (amplification conditions).

**Table 1:**
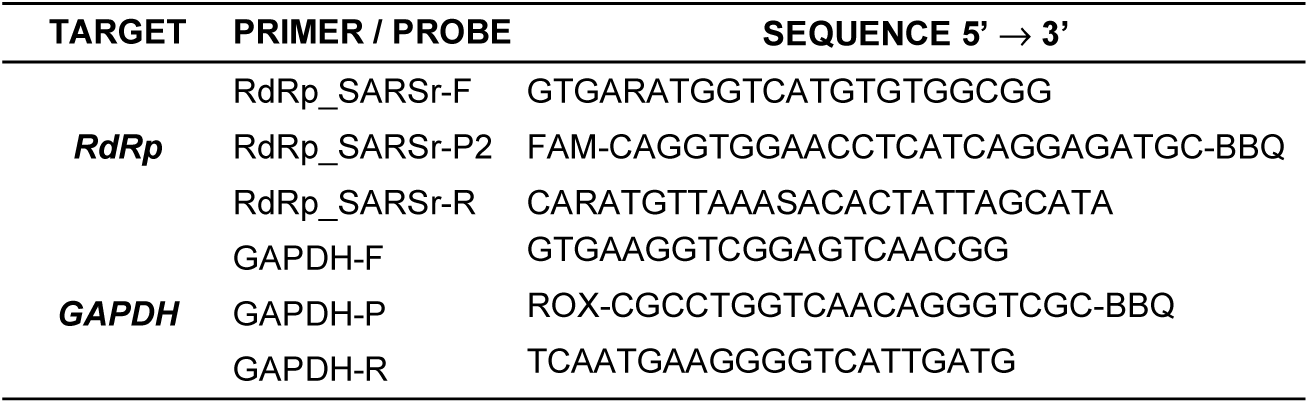
Target genes, oligonucleotides and probes used in the RT-qPCR reactions. The targets for amplification were RNA dependent RNA polymerase (*RdRp*) specific for SARS-CoV-2 and the Glyceraldehyde-3-Phosphate Dehydrogenase (*GAPDH*), a human constitutive gene. The sample quality, the RNA extraction and amplifications performances were evaluated in a single multiplex reaction using GAPDH as internal control.

**Table 2:** Conditions of RT-qPCR multiplex reactions for SARS-CoV-2 and GAPDH using Capital™ RT-qPCR Probe Mix 4X (Biotechrabbit).

| REAGENTS and CONCENTRATIONS | VOLUME (μL) |
| --- | --- |
| RdRp_SARSr-F (10 μM) | 0.8 |
| RdRp_SARSr-P2 (10 μM) | 0.8 |
| RdRp_SARSr-R (10 μM) | 0.4 |
| GAPDH-F (2.5 μM) | 0.5 |
| GAPDH-P (2.5 μM) | 0.5 |
| GAPDH-R (1.25 μM) | 0.4 |
| RTase with RNase inhibitor | 1.0 |
| qPCR PROBE MIX | 5.0 |
| Nuclease Free Water | 5.6 |
| **FINAL VOLUME** | **15.0** |

**Table 3:**
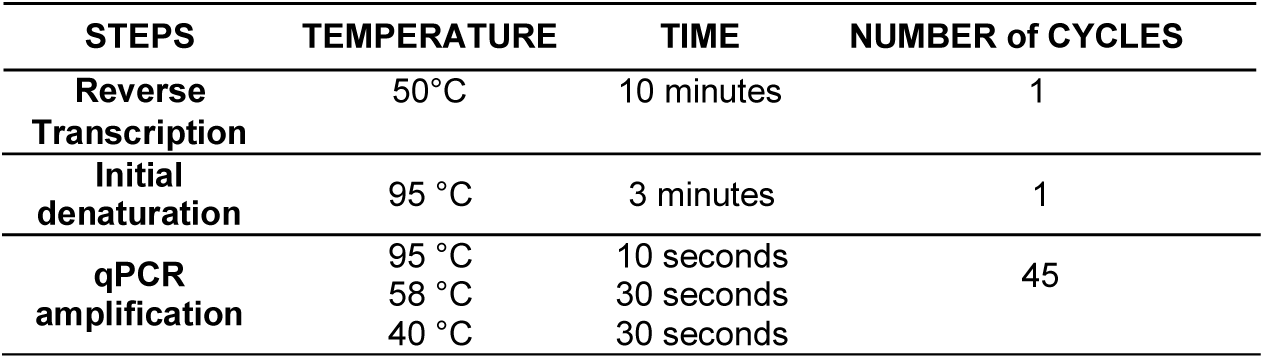
Conditions of RT-qPCR multiplex amplification for SARS-CoV-2 and GAPDH using Capital™ RT-qPCR Probe Mix 4X (Biotechrabbit).

#### 2.4.3. RT-LAMP REACTION

The RT-LAMP reactions were performed according to Lamb et al. (2020) [8] using WarmStart Colorimetric LAMP 2X Master Mix, containing a pH indicator which allows the colorimetric visualization. The robustness was tested from standard primers concentration and final volume of reaction. The concentrations of reagents and the reactions conditions were summarized in table 4.

**Table 4:**
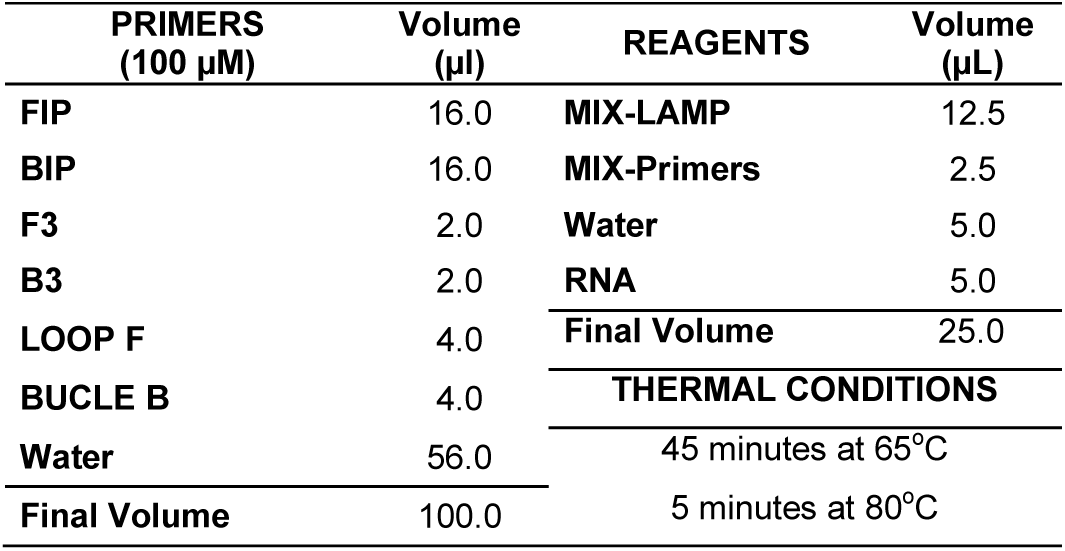
Conditions of RT-LAMP reactions to detect SARS-CoV-2, according to Lamb et al. (2020). The primers’ names were the same on the original publication.

### 2.5. STATISTICAL ANALYSIS

Data analysis was performed using the Stata v16.1 statistical package (Stata Corporation, College Station, Texas, USA); point estimators and 95% confidence intervals of the clinical-epidemiological characteristics of the people evaluated were calculated. The values of the diagnostic performance measures of RT-LAMP in comparison with RT-qPCR were calculated; considering: sensitivity, specificity, positive and negative predictive value, positive and negative likelihood ratio, area under the ROC curve, Matthews Correlation Coefficient and F1-Score. The degree of concordance between the results of both tests was determined, as well as the agreement using Cohen’s Kappa index. These analyzes were carried out for all evaluated cases, as well as in a stratified way according to week of illness. The relationship between time of symptoms and Ct values was established using Pearson’s correlation coefficient.

## 3. RESULTS

### 3.1. LABORATORY STANDARDIZATION

The limit of detection for SARS-CoV-2 by RT-LAMP was consistent only with those with Ct values < 30 in the RT-qPCR reactions (standard curve presented into figure 1, panel A, and RT-LAMP performance reaction, panel B) and RT-LAMP in table 5. This means that the RT-LAMP test was efficient to detect up to 1000 copies/µL of the target gene. In the robustness experiments, high reactions performances were obtained with half of primers concentrations (0.5P) and with 20 µL of final volume (0.8V from final volume of standard reaction).

**Table 5:** Comparison of limit of detection between RT-qPCR and RT-LAMP reactions to detect SARS-CoV-2.

| SERIAL DILUTION | $10^{-1}$ | $10^{-2}$ | $10^{-3}$ | $10^{-4}$ | $10^{-5}$ | $10^{-6}$ | $10^{-7}$ |
| --- | --- | --- | --- | --- | --- | --- | --- |
| CONCENTRATION<br>(number of copies/μL) | $10^7$ | $10^6$ | $10^5$ | $10^4$ | $10^3$ | $10^2$ | $10^1$ |
| Ct VALUES (RT-qPCR) | 13.59 | 16.70 | 20.37 | 25.04 | 29.17 | 35.12 | - |
| COLOR CHANGE (RT-LAMP) | Yes | Yes | Yes | Yes | Yes | No | No |

**Figure 1:**
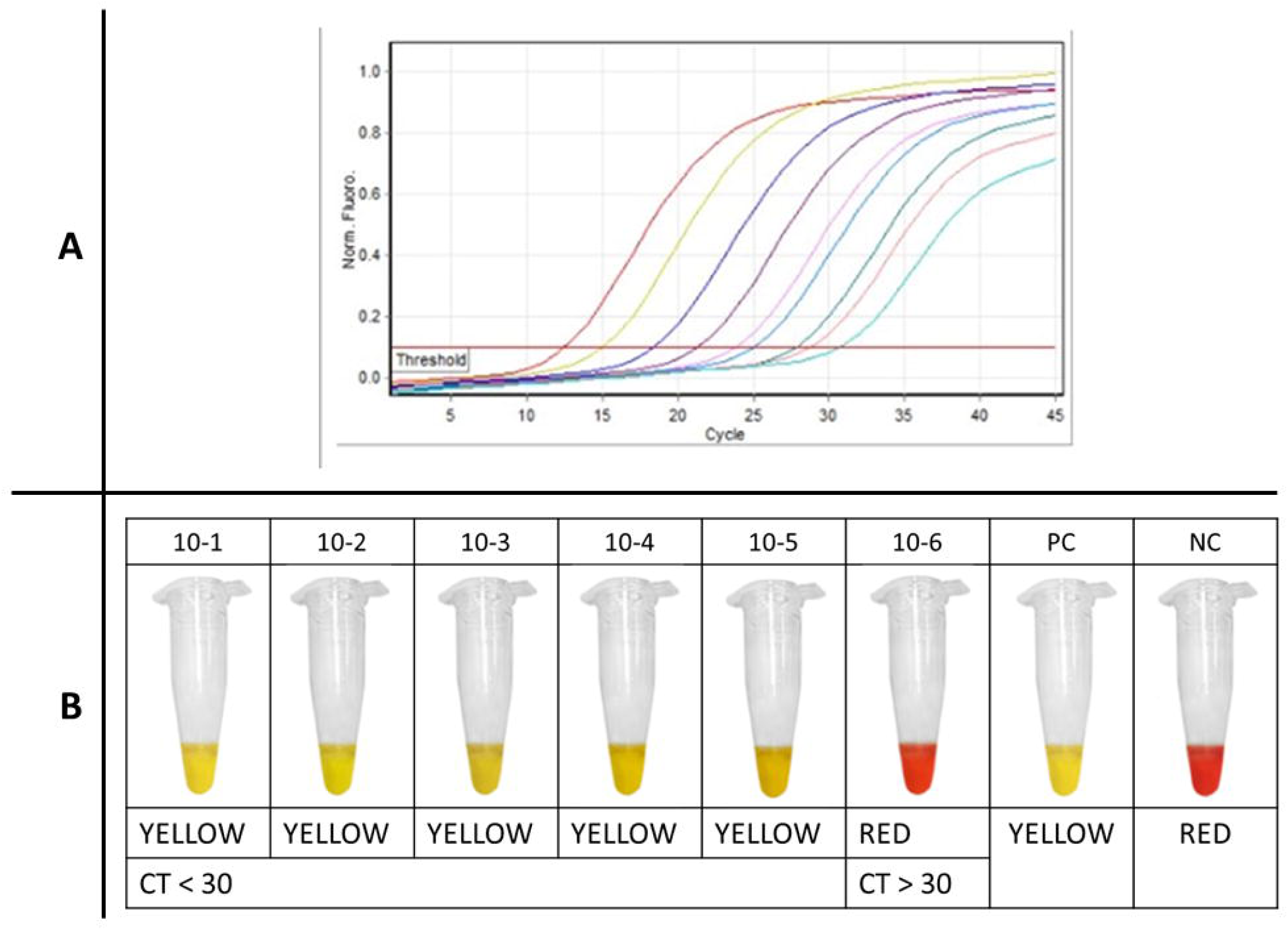
Standard curve of RT-qPCR (panel A) reactions and limit of detection by RT-LAMP (panel B) in two molecular methods to detect SARS-CoV-2.

The “cross-reaction analysis” performed *in silico* identified a very low-similarity degrees between the primers alignment and reference sequences of HCoV NL-63, HKU1, OC43, 229E, SARS-CoV-1, MERS and HECoV (figure 2: panel A refers to F3 primer alignment – forward; panel B refers to B3 primer alignment – reverse). These data, would indicate the absence of amplification of other HCoV, if they to be present in the sample. The yellow columns correspond to conserved regions. In addition, when these same primers were aligned with 194 Peruvian strains made available on GISAID initiative, there was none exclusion of conserved regions, exhibiting a high-similarity and specificity, which may be designated as absence of concomitant detection of other HCoV non-SARS-CoV-2.

**Figure 2:**
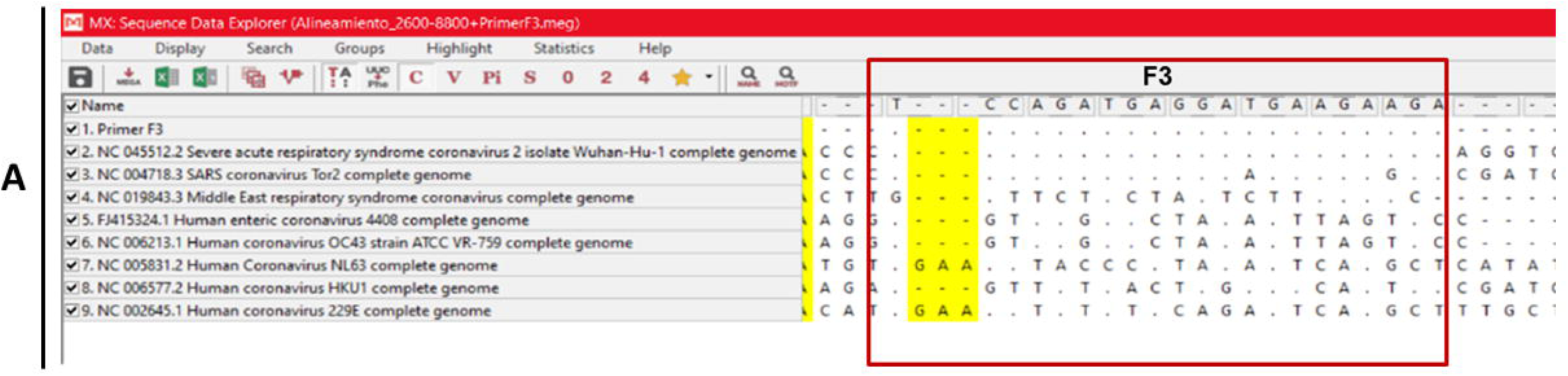
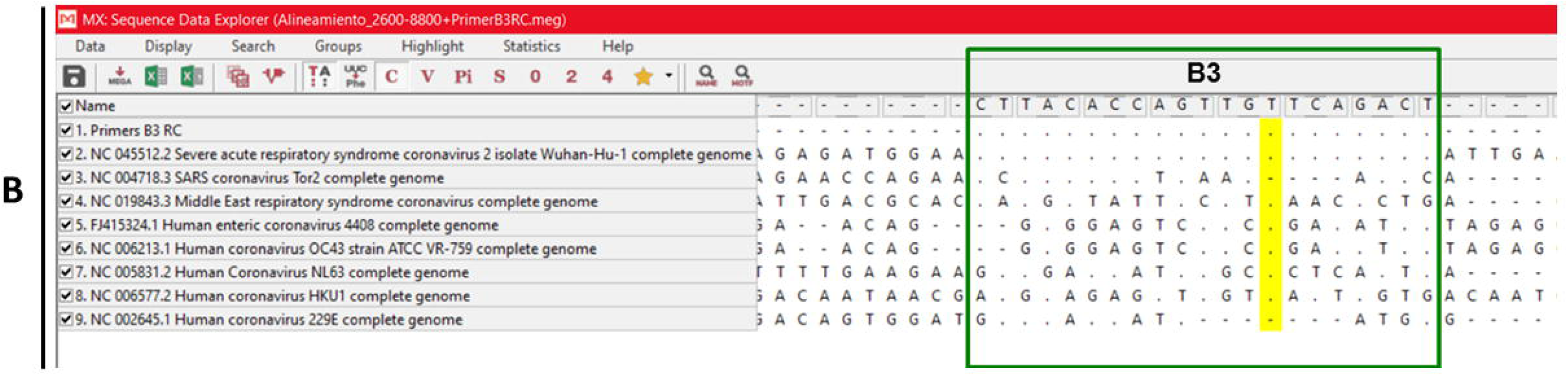
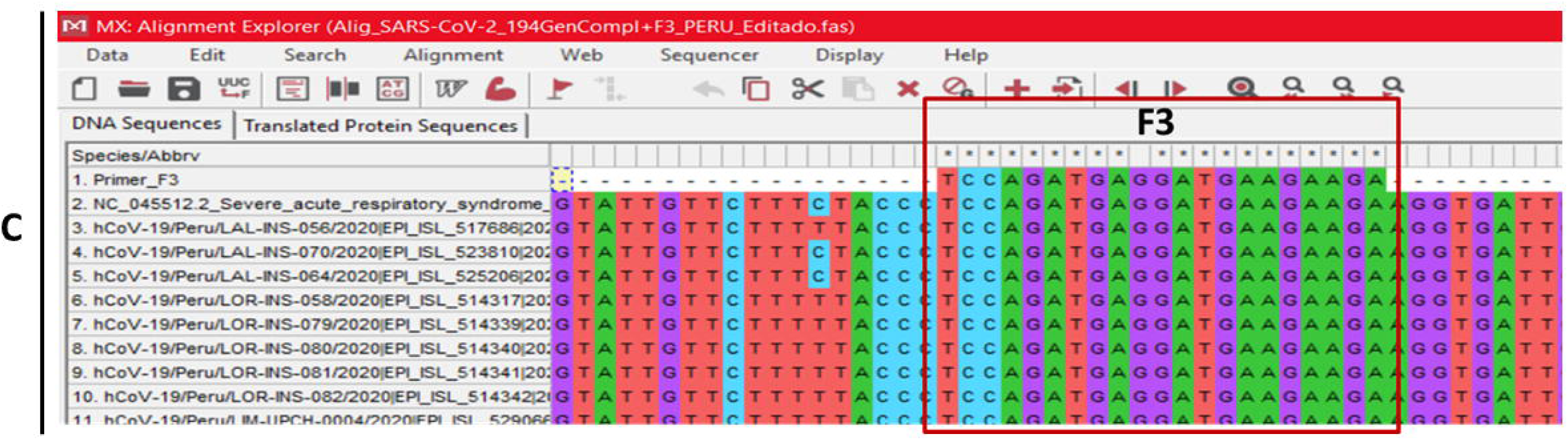
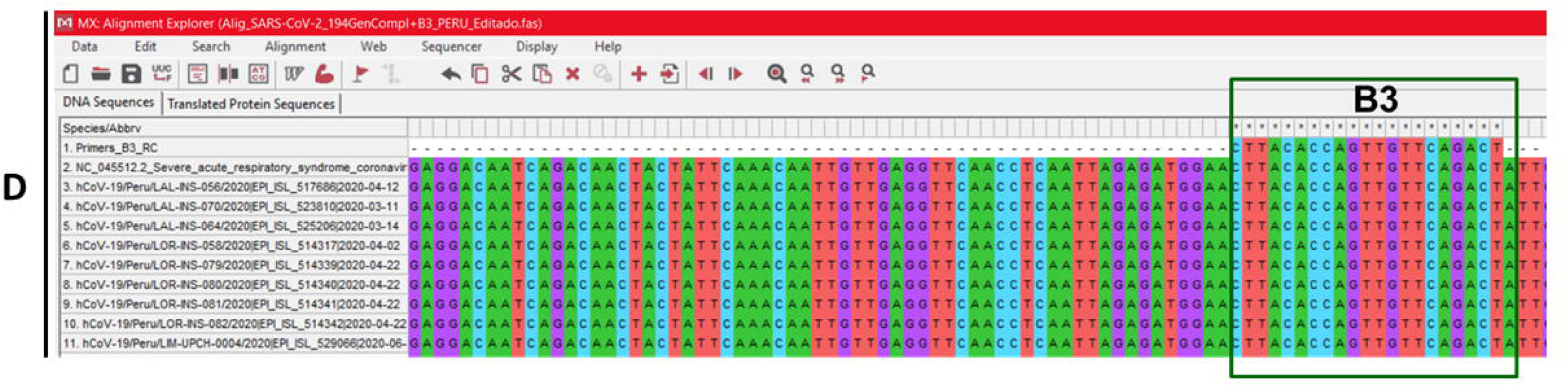
Multiple sequence alignment between RT-LAMP external primers F3 and B3 (Lamb et al., 2020) and reference sequences of all known human coronaviruses and all SARS-CoV-2 Peruvian strains made available on GISAID initiative. The alignment was conducted in ClustalW using MEGA. The primers sequences (panel A – F3, panel B – B3) were aligned with all reference sequences of known HCoV (NC_005831.2, HCoV-NL63; NC_002645.1, HCoV-229E; NC_006213.1, HCoV-OC43 strain ATCC VR-759; NC_006577.2, HCoV-HKU1; NC_004718.3, SARS-CoV-1; NC_019843.3, MERS; FJ415324.1, HECoV-4408 and NC_045512.2, SARS-CoV-2 isolate Wuhan-Hu-1) and all 194 SARS-CoV-2 Peruvian strains (panel C – F3, panel D – B3). The yellow columns, on the panels A and B, and asterisks, on the panels C and D, represent conserved regions into *nsp3* gene fragment between the all known HCoV and all SARS-CoV-2 Peruvian strains complete genome, respectively.

The positivity obtained for each method, RT-qPCR and RT-LAMP, is presented in table 6. The values of Cohen’s kappa index comparing the diagnostic performance between both methods indicated a nearly perfect agreement between them, with the best agreement on the onset of symptoms.

**Table 6:**
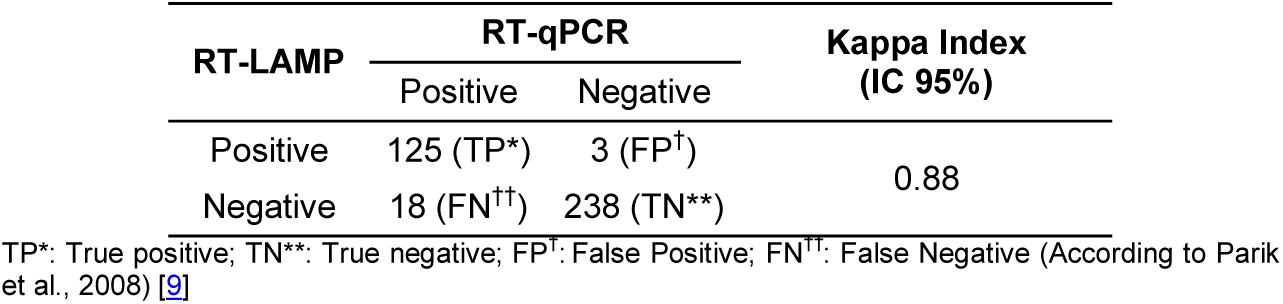
Results obtained in the laboratory standardization for performance diagnostic comparison between RT-LAMP and RT-qPCR in the SARS-CoV-2 molecular detection.

| RT-LAMP | RT-qPCR |  | Kappa Index<br>(IC 95%) |
| --- | --- | --- | --- |
|  | Positive | Negative |  |
| Positive | 125 (TP*) | 3 (FP<sup>†</sup>) | 0.88 |
| Negative | 18 (FN<sup>††</sup>) | 238 (TN**) |  |

TP\*: True positive; TN\*\*: True negative; FP<sup>†</sup>: False Positive; FN<sup>††</sup>: False Negative (According to Parik et al., 2008) [9]

### 3.2. CLINICAL ASSESSMENT

The study population was composed by 51.7% (n = 198) women and 48.3% (n = 185) men, being young adults the most frequent age group (n = 236, 61.6%). The most common symptoms were cough (n = 268, 70.0%) and pharyngeal pain (n = 262, 68.4%). Regarding time of symptom onset, the average was 7.1 ± 3.3 days, and 56.3% belong to the first week after symptom onset patients (group 1) and 43.7% belong to the second week after symptom onset patients (group 2). One case was excluded due to memory bias.

We determined 37.3% positive samples by RT-qPCR and 33.7% by RT-LAMP (table 6). Among the 143 positive results by RT-qPCR, only 20 clinical samples had discordant results with RT-LAMP, 17 were false negatives and 3 were false positives. In group 1, the Ct was between 31.00 and 36.46, with a median of 34.43 (IQR: 34.2, 35.56). In group 2, the Ct values were higher than 37. The true positive data presented significant concordance (p <0.001) between both tests (Kappa index: 88.6; 95% CI between 83.8 and 93.5); being in group 1, 70 of 76 positive cases (Kappa index: 91.8; 95% CI between 86.2 and 97.4), while in group 2, 56 of 67 cases (Kappa index: 84.7; 95% CI between 76.4 and 93.0]) (table 7).

**Table 6:**
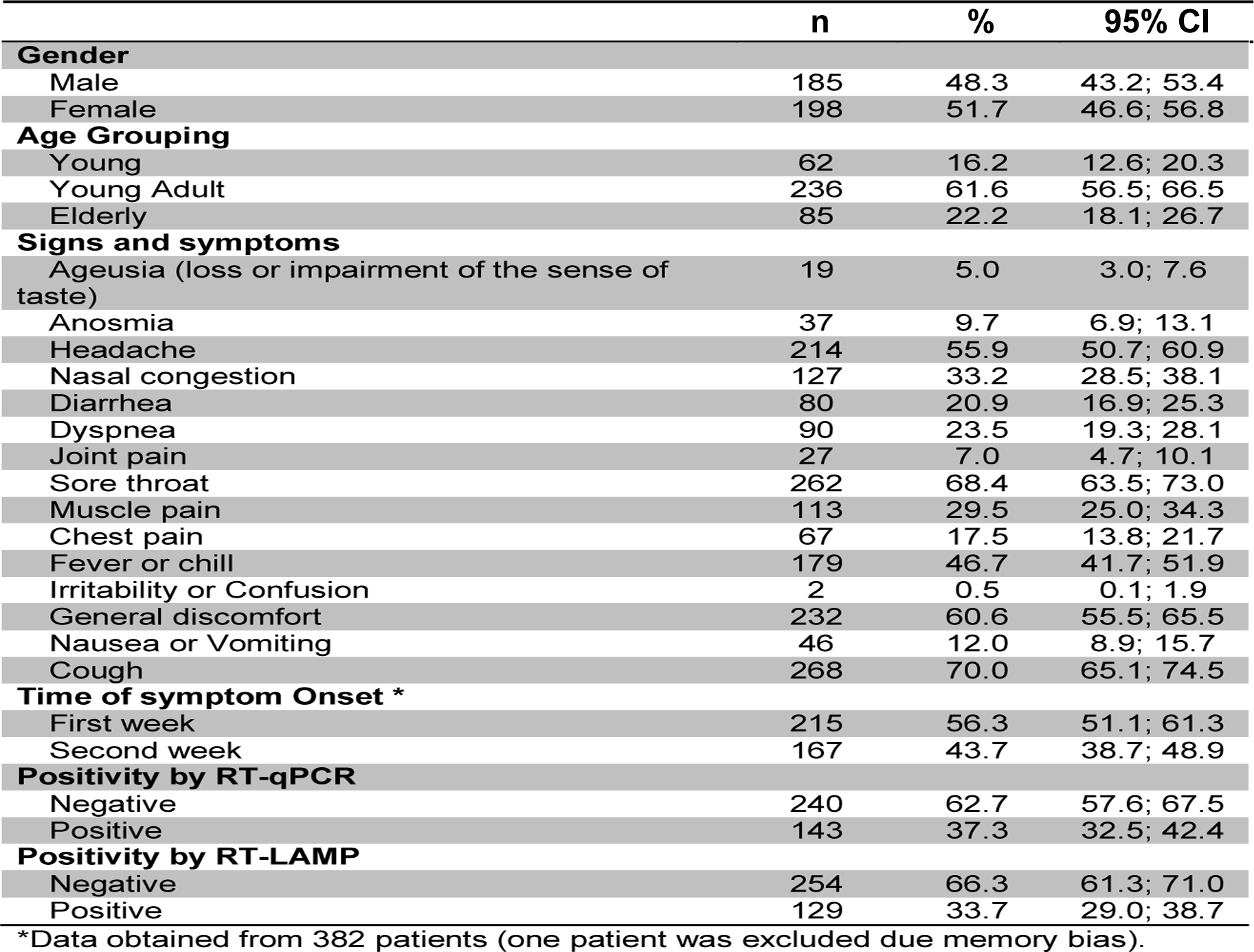
Clinical and epidemiological data related to the study population (gender, age grouping, signs and symptoms, time of illness onset, positivity by RT-qPCR and by RT-LAMP.

|  |  |  |  |
| --- | --- | --- | --- |
| **Gender** |  |  |  |
| Male | 185 | 48.3 | 43.2; 53.4 |
| Female | 198 | 51.7 | 46.6; 56.8 |
| **Age Grouping** |  |  |  |
| Young | 62 | 16.2 | 12.6; 20.3 |
| Young Adult | 236 | 61.6 | 56.5; 66.5 |
| Elderly | 85 | 22.2 | 18.1; 26.7 |
| **Signs and symptoms** |  |  |  |
| Ageusia (loss or impairment of the sense of taste) | 19 | 5.0 | 3.0; 7.6 |
| Anosmia | 37 | 9.7 | 6.9; 13.1 |
| Headache | 214 | 55.9 | 50.7; 60.9 |
| Nasal congestion | 127 | 33.2 | 28.5; 38.1 |
| Diarrhea | 80 | 20.9 | 16.9; 25.3 |
| Dyspnea | 90 | 23.5 | 19.3; 28.1 |
| Joint pain | 27 | 7.0 | 4.7; 10.1 |
| Sore throat | 262 | 68.4 | 63.5; 73.0 |
| Muscle pain | 113 | 29.5 | 25.0; 34.3 |
| Chest pain | 67 | 17.5 | 13.8; 21.7 |
| Fever or chill | 179 | 46.7 | 41.7; 51.9 |
| Irritability or Confusion | 2 | 0.5 | 0.1; 1.9 |
| General discomfort | 232 | 60.6 | 55.5; 65.5 |
| Nausea or Vomiting | 46 | 12.0 | 8.9; 15.7 |
| Cough | 268 | 70.0 | 65.1; 74.5 |
| **Time of symptom Onset \*** |  |  |  |
| First week | 215 | 56.3 | 51.1; 61.3 |
| Second week | 167 | 43.7 | 38.7; 48.9 |
| **Positivity by RT-qPCR** |  |  |  |
| Negative | 240 | 62.7 | 57.6; 67.5 |
| Positive | 143 | 37.3 | 32.5; 42.4 |
| **Positivity by RT-LAMP** |  |  |  |
| Negative | 254 | 66.3 | 61.3; 71.0 |
| Positive | 129 | 33.7 | 29.0; 38.7 |

\*Data obtained from 382 patients (one patient was excluded due memory bias).

**Table 7:** Results from clinical validation, comparing diagnostic performance between RT-LAMP and RT-qPCR for SARS-CoV-2 molecular detection. This data was used to calculate the sensitivity, specificity, predictive positive (PPV) and predictive negative (PNV) values.

| RT-LAMP | RT-qPCR |  | Kappa Index<br>(IC 95%) | Concordance<br>% | p-value |
| --- | --- | --- | --- | --- | --- |
|  | Positive | Negative |  |  |  |
| Overall |  |  |  |  |  |
| Positive | 126 (TP*) | 3 (FP<sup>†</sup>) | 0.88 (0.83; 0.93) | 94.8 | <0.001 |
| Negative | 17 (FN<sup>††</sup>) | 237 (TN<sup>**</sup>) |  |  |  |
|  | Positive | Negative |  |  |  |
| First week of symptoms |  |  |  |  |  |
| Positive | 70 (TP*) | 2 (FP<sup>†</sup>) | 0.91 (0.86; 0.97) | 96.3 | <0.001 |
| Negative | 6 (FN<sup>††</sup>) | 137 (TN**) |  |  |  |
| Second week of symptoms |  |  |  |  |  |
| Positive | 56 (TP*) | 1 (FP<sup>†</sup>) | 0.84 (0.76; 0.93) | 92.8 | <0.001 |
| Negative | 11 (FN<sup>††</sup>) | 99 (TN**) |  |  |  |

TP\*: True positive; TN\*\*: True negative; FP<sup>†</sup>: False Positive; FN<sup>††</sup>: False Negative (According to Parik et al., 2008) [9].

### 3.3. DIAGNOSTIC PERFORMANCE of RT-LAMP

The sensitivity was 88.1% (95% CI: 81.6; 92.9) and the specificity, 98.8% (95% CI: 96.4; 99.7). These values presented variations when a stratified evaluation was made by time of symptom onset; where the sensitivity reaches 92.1% in the first week, while for the second it drops to 86.6%. However, the specificity showed minimal variation, with 98.6% and 99.0% for the first and second week respectively (table 8).

**Table 8:** Laboratory and clinical performance of RT-LAMP using RT-qPCR as reference test.

| PARAMETERS | LABORATORY<br>STANDARDIZATION |  | CLINICAL ASSESSMENT |  |  |  |  |  |
| --- | --- | --- | --- | --- | --- | --- | --- | --- |
|  |  |  | Overall |  | First week of<br>symptoms |  | Second week of<br>symptoms |  |
|  | % | 95% CI | % | 95% CI | % | 95% CI | % | 95% CI |
| **Sensitivity\*** | 87.4 | 80.8; 92.4 | 88.1 | 81.6; 92.9 | 92.1 | 83.6; 97.0 | 86.6 | 72.5; 91.5 |
| **Specificity\*\*** | 98.8 | 96.4; 99.7 | 98.8 | 96.4; 99.7 | 98.6 | 94.9; 99.8 | 99.0 | 94.6; 100 |
| **Positive Predictive Value†** | 97.7 | 93.3; 99.5 | 97.7 | 93.4; 99.5 | 97.2 | 90.3; 99.7 | 98.2 | 90.6; 100 |
| **Negative Predictive Value‡** | 93.0 | 89.1; 95.8 | 93.3 | 89.5; 96.1 | 95.8 | 91.0; 98.4 | 90.0 | 82.8; 94.9 |
| **Accuracy** | 94.5 | 91.8; 96.6 | 94.8 | 92.1; 96.8 | 96.3 | 92.8; 98.4 | 92.8 | 87.8; 96.2 |
| **Area Under curve** | 93.1 | 90.3; 95.9 | 93.4 | 90.7; 96.2 | 95.3 | 92.1; 98.5 | 91.3 | 86.7; 95.9 |
| **Matthews Correlation Coefficient** | 88.4 | ---- | 88.9 | ---- | 91.8 | ---- | 85.4 | ---- |
| **F1 Score** | 92.3 | ---- | 92.6 | ---- | 94.6 | ---- | 90.3 | ---- |

\*Sensitivity [(TP)/(TP+FN)]; \*\*Specificity [(TN)/(TN+FP)]; †PPV [(TP)/(TP+FP)]; ‡NPV [(TN)/(TN+FN)] (According to Parik et al., 2008) \[9\].

**Table 10.** Correlation between Ct values and time of illness onset.

|  | Overall |  | First week of symptoms |  | Second week of symptoms |  | Rho† | P-Value |
| --- | --- | --- | --- | --- | --- | --- | --- | --- |
|  | Positive | Median (RIQ) | Positive | Median (RIQ) | Positive | Median (RIQ) |  |  |
| **RT-qPCR** | 143 | 29.4 (7.8) | 76 | 27.6 (8.0) | 67 | 30.5 (7.3) | 0.326 | 0.001 |
| **RT-LAMP** | 126 | 28.4 (6.9) | 70 | 27.4 (7.4) | 56 | 29.9 (6.7) | 0.350 | 0.001 |

†Spearman's Correlation Coefficient.

The positive predictive value was 97.7%, which showed an increase according to the time of symptom onset, from 97.2%, in the first week, to 98.2%, for the second. It was identified that the overall positive likelihood ratio of the test was 70.5, taking values of 64.0 and 83.6 in the first and second week respectively. Meanwhile, the negative likelihood ratio was 0.12, with values of 0.08 in the first week, and 0.17 for the second. The accuracy tends to show lower results as the time of illness onset progresses.

### 3.4. RELATIONSHIP BETWEEN Ct VALUES AND DINAMICS OF VIRAL INFECTION

The overall median Ct value was 29.4 (7.8), 27.6 (8.0) for the first week of symptom onset and 30.5 (7.3) for the second week. The overall median Ct values obtained by RT-qPCR from all positive samples by RT-LAMP was 28.4 (7.0), 27.4 (7.4) in the first week, and 29.9 (6.7) in the second. A non-linear trend was found for higher Ct values as there was a longer time of symptom onset. A direct relation of 32.6% was identified between the Ct values of the positive cases detected by RT-qPCR with the time of symptom onset (p = 0.001). Meanwhile, for the positive cases according to RT-LAMP, the correlation between Ct values and time of symptom onset was 35.0% (p = 0.001) (figure 3, table 9).

**Figure 3:**
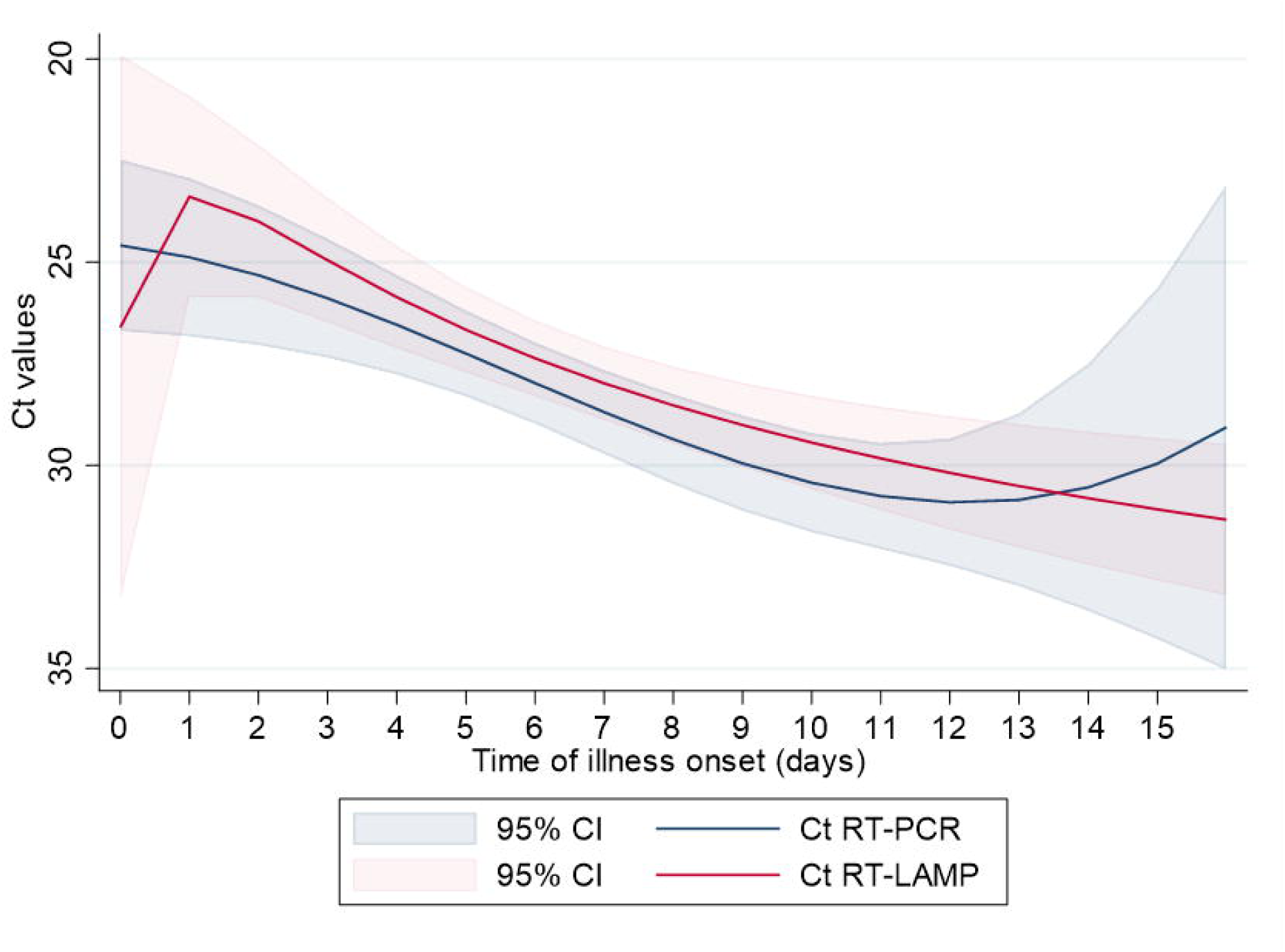
Distribution of Ct values obtained by RT-qPCR (reference test) using positivity data obtained in the both methods, RT-qPCR and RT-LAMP. The results indicated that the Ct values increase with the course of the disease, suggesting a decrease of viral load.

## 4. DISCUSSION

In Peru, the first measure adopted to contain the virus dissemination was the quarantine that endured between March 16^th^ and June 30^th^. On this long and difficult time, many diagnostic strategies were implemented and until now, almost 300,000 samples have been processed by RT-qPCR only in the COVID-19 Emergency Laboratory from Instituto Nacional de Salud [10]. Even though other molecular biology laboratories have been implemented in different regions of the country, this strategy have not been enough to contain the virus dissemination in our country. Peru is the sixth country of the world in total number of COVID-19 positive cases and the first in the mortality (96 deceased for every 100,000 inhabitants) [11].

On the other hand, the sample transport logistics between collection point and processing remains as a problem to overcome. In this sense, the molecular test available at the healthcare unit should be a good strategy to detect on time and control the SARS-CoV-2 transmissibility. To select the best diagnostic strategy, some challenges must be considered. Additionally, It is essential to have clarity about the purpose, regulatory approval, diagnostic accuracy under ideal conditions, data on the diagnostic accuracy in clinical practice and finally, the test’s performance used in routine use publicly available [12]. So, the method chosen must no require complex equipment or specialized human resources, must be fast producing results in short time and must be comparable to RT-qPCR, the gold standard molecular method recommended by WHO. Considering all these points, the RT-LAMP can be a feasible alternative for all these requirements.

Considering the geographic and economic structure of Peru that implies directly in the logistic transport and epidemiological conditions of several infectious diseases, the Ministry of Health and INS have gradually produced and implemented molecular diagnostic tests based on RT-LAMP method for cholera, febrile disease caused by arboviruses Zika, and Dengue. Since other researchers, during this pandemic, already described several RT-LAMP for SARS-CoV detection [7, 8, 13, 14, 15, 16, 17, 18 and 19], the INS Peruvian researchers’ team selected the protocol described Lamb et al (2020) to compare its performance diagnostic in comparison with RT-qPCR. This protocol is based in a fast-colorimetric reaction and can provide results in less than 60 minutes after RNA extraction.

We compared the diagnostic performance of this specific protocol in two steps of quality verification. The first step was performed as laboratory standardization and, the second one, as clinical validation. In these two phases, 767 clinical samples were processed and the results indicated that this protocol have similar diagnostic performance when compared to RT-qPCR. The limit of detection of this method was 1,000 copies/µL (table 5 and figure 1), ten times lower than RT-qPCR standardized and implemented in the molecular diagnostic routine by INS. However, this difference should be associated to the target gene for the methods to be different and to be in different *ORFs*. The primers for RT-LAMP were designed to align in the *ORF1a* region, to detect a SARS-CoV-2 *nsp3* gene fragment and the primers for RT-qPCR, into the *ORF1b*, for in *RdRp* gene fragment. Considering the CoV replication, many subgenomic RNA are generated in different quantities and this particular characteristic should be considered in the molecular test using different target gene [20]. From these replication characteristics of Coronaviridae family, the WHO has suggested that the diagnosis should be conducted using primers for Nucleocapsid (*N*) gene or for *ORF1ab* genes. Even so, since the *ORF1ab* represents 2/3 of all genome (reference sequence NC_045512.2), it should be considered that the genes located on the 5’ genome has less copy during replication cycle. Therefore, the *nsp3* gene may have a lower amount of RNA during replication when compared to the amount of RNA for the *RdRp* gene, which would justify the lower sensitivity of the RT-LAMP test. To overcome these difficulties, we designed new set of primers for others genome regions, especially for *RdRp*, to properly compare the diagnostic performance considering the same genomic region.

We also showed by *in silico* analysis that the set of primers used for RT-LAMP was really specific to detect the SARS-CoV-2 Peruvian strains and did not present cross-reaction with others HCoV in molecular test (figure 2, panels A and B). We know that this point was a limitation of our study because this analysis should be done in vitro using clinical samples. Furthermore, the INS does not have positive clinical samples for other HCoV. Due to the need to quickly evaluate the performance of this diagnostic method and finally start transferring this technology to the points of attention, the alternative of verifying the occurrence of cross reaction measured by in silico analysis was the most appropriate and scientifically feasible at the moment. The perfect identity in the region of primers alignment F3 and B3 with all available SARS-CoV-2 Peruvian strains (figure 2, panels C and D) also indicated specific detection and almost none probability of false negative results due primers specificity.

The robustness evaluation of this RT-LAMP protocol considered variables as primers concentration and final volume of reaction. This strategy focused the fact of the reactions will be performed by people that does not present routine contact with molecular biology techniques. Since the reactions performance was not compromised using half of primers concentrations and eighty percent of final volume of reaction, technical errors that may be made during small volume pipetting.

The RT-LAMP presented a high sensitivity and specificity in the both steps of quality verification (87.4% and 98.7%, 88.1% and 98.8%, available in table 8 obtained by results presented in tables 6 and 7 in the laboratory standardization and in the clinical validation, respectively). These results were similar to those reported by Hu et al. (2020) [21] (88.57% and 98.98%, respectively) and lower than described by Jiang et al. (2020) [8] (91.4% and 99.5%, respectively) and Kitagawa et al. (2020) [22] (100% and 97.6%, respectively). These differences could be associated to the Ct values used to establish positivity by RT-qPCR. Furthermore, only the positive samples that presented Ct values > 30 disagreed with those obtained by RT-LAMP. So, our results indicated 100% specificity and sensitivity because Ct > 30 exceeds the minimum number of copies that represents the limit of detection of this protocol. In addition, the Kappa index about 0.9 showed a virtually perfect agreement between these tests, indicating that this RT-LAMP protocol can be used as alternative method of COVID-19 molecular diagnosis at healthcare centers.

The area under the curve of the RT-LAMP test was 93.4% for the clinical assessment. We did not find any article that has reported this aspect for the RT-LAMP. However, as it is very close to 100%, it reflects that RT-LAMP can be useful enough to identify infected patients in the active transmission phase.

It was found that the RT-LAMP test, when giving a Positive Predictive Value 97.7%, in a similar way to that reported by Jiang et al. (2020) [7], PPV: 97.7%), and much higher than mentioned by Hu et al. (2020) [21] (PPV: 91.18%), the latter evaluated 329 nasal and pharyngeal swabs from a cohort of 129 COVID-19 suspects and serial upper respiratory tract samples from asymptomatic carriers, unlike our study in which only samples of symptomatic cases. Similarly, when giving a negative result, the RT-LAMP succeeded in 93.3% of the cases in identifying a person without SARS-CoV-2 infection, which although it is somewhat lower than that reported by Jiang et al. (2020) [7], who found a Negative Predictive Value (NPV) of 98.1%. This difference also could be associated to prevalence obtained in each study.

The degree of agreement or concordance in the identification of SARS-CoV-2 between RT-qPCR and RT-LAMP at clinical assessment was 94.8%, which indicated that there was a great concordance degree between the tests, similar to that found in other studies such as the one by Lu et al. (2020) [16], and Kitagawa et al. (2020) [22], where it was always greater than 90%. In contrast, we found 20 discordant results between RT-LAMP and RT-qPCR in the clinical assessment, 17 false negatives and 3 false positive; Jiang et al. (2020) [7] also found 5 discordant results, 4 false negatives and 1 false positive. Kitagawa et al. (2020) [22] reported only 2 discordant, which were false positives. Hu et al. (2020) [21] also identified 4 discordant samples, theoretically false positives; however, these were confirmed as SARS-CoV-2 positive through a genetic sequencing test.

When evaluating the performance of the RT-LAMP by time of symptom onset, we found that the sensitivity and the Negative Predictive Value were higher in the first week, and although the Positive Predictive Value and the specificity showed an increase towards the second week, although this increase was not significant. We did not find any article that evaluates the performance of RT-LAMP by time of symptom onset, but RT-qPCR shows greater performance in the first week of symptoms; these findings could be verified with the area under the curve, which from being 95.3% in first week it is reduced to 91.3% at second week of the days onset of symptoms.

Within the clinical limitations, it should be mentioned that the RT-LAMP test was only evaluated in symptomatic cases. However, the purpose of this study was to evaluate a simple, sensitive, specific and robust, low-cost diagnostic method to be implemented in healthcare units.

Finally, our data allow us to conclude that the RT-LAMP protocol implemented by INS should be the convenient alternative for SARS-CoV-2 detection directly at the healthcare centers in this moment. This strategy can provide appropriate prevention and control measures in all provinces and for decreasing the number of severe and non-severe cases of COVID-19.

## Data Availability

All data are fully available without restriction

## 5. ACKNOWLEGMENTS

We thank Pan American Health Organization (PAHO) for providing us the reagents and to stablish collaboration to conduct the experiment validations. Our recognition to all the workers of Laboratorio de Microbiologia y Biomedicina of INS and all people from others institutions involved in obtaining, handling and processing the samples, in special to Jairo Mendez (PAHO), Rapid Response Team (CDC/INS), Lely Solari, Faviola Valdivia, Helen Horna, Gabriel de Lucio, Yanina Zarate, Iris Pompa, Isidro Antipupa,, Jhon Mayo, Carina Mantari, Kathia Tarqui, Romeo Pomari, Eduardo Juscamayta, Paquita García, Miryam Palomino, Pamela Rios, Priscila Lope, Johana Balbuena, Victor Jiménez, Yolanda Angulo, Yuli Barrios, Paul Pachas, Noemi Flores, and Ana Zeppilli.

